# Investigation of the correlation of adropin with anthropological and psychological factors in schizophrenia: preliminary evidence from a case-control study

**DOI:** 10.64898/2026.02.20.26346678

**Authors:** Yuko Nishida, Ryusei Nishi, Takamasa Fukumoto, Ei’ichi Iizasa, Yoshiaki Nishida, Akihiro Asakawa

## Abstract

**Background and Hypothesis:** Schizophrenia is a disease characterized by various symptoms and has severe lifelong impacts on patients and their families. Despite various hypotheses and associated studies, the key mechanism in schizophrenia is not fully elucidated. In the present study, we focused on adropin, a peptide regulating energy metabolism, antioxidation, and neuroprotection.

**Study Design:** In both the group of healthy volunteers (HV) and the group of patients with some schizophrenia spectrum and other psychotic disorders (SZ), we evaluated adropin along with other variables such as anthropological factors, psychological well-being indicators, and laboratory test results.

**Study Results:** The adropin levels in SZ were not significantly different from those in HV. Correlation analysis indicated five significant correlations beyond various natural correlations arising from fundamental proportional relationships and multifaceted psychological well-being indicators: (1) adropin versus right handgrip strength in the SZ group (τ = –0.82, *P* = 0.066); (2) adropin versus selenium in the total group (τ = 0.44, *P* = 0.053); (3) ferritin versus perceived stress in the total group (τ = –0.44, *P* = 0.053); (4) right versus left handgrip strength in the total group (τ = 0.70, *P* = 0.001) and in the SZ group (τ = 0.82, *P* = 0.075); and (5) selenium versus state anxiety in the total group (τ = 0.44, *P* = 0.053) and the SZ group (τ = 0.84, *P* = 0.066).

**Conclusions:** The present study provides a foundation for future studies and sheds light on the role of adropin in schizophrenia.

## Introduction

Schizophrenia, a disease characterized by various symptoms, including positive, negative, and disorganization symptoms,^1^ is a severe and complex mental disorder that affects society. Patients with schizophrenia not only suffer from psychiatric symptoms but also from severe functional impairments, which antipsychotics cannot improve.^2^ The social and economic burden on family caregivers reduces the quality of life and overwhelms them emotionally.^3^ The prevalence has traditionally been estimated at 1 out of 100, which is considered relatively high considering the severity of symptoms and their lifelong impacts. In Japan, the total cost in 2008 was estimated to be 23.8 billion USD,^4^ which accounted for approximately 10 percent of the national social security costs.

The main hypothesis of schizophrenia is dysfunction in neurotransmission, and there is substantial evidence for the two main neurotransmitters, dopamine and glutamate.^1^ Furthermore, other promising targets have been studied, such as muscarinic receptors (M_1_ and M_4_) and glycine transporter 1.^5^ Despite various hypotheses and associated studies, the key mechanism in schizophrenia is not fully elucidated. Recently, schizophrenia has been studied from the perspective of brain energy metabolism. Antipsychotics such as haloperidol and risperidone impair complex I activity in mitochondria, hindering cellular respiration and thus leading to adenosine triphosphate depletion or increasing reactive oxygen species.^6–8^

Various peptides and neurons are involved in brain metabolism. Recently, a 76-amino acid polypeptide of adropin was identified.^9^ Adropin is encoded by the energy homeostasis-associated gene located at chromosome 9p13.3, and is widely expressed, including in the brain. Adropin might play a crucial role in energy metabolism,^10^ as evidenced by its activation of the phosphatidylinositol-3 kinase/protein kinase B and extracellular signal-regulated kinase pathway, which is important in cancer,^11^ or by protecting endothelial cells through the expression of endothelial nitric oxide synthase, which facilitates nitric oxide production.^12,13^

Considering schizophrenia from the perspectives of energy metabolism and the role of adropin therein, it might be natural to hypothesize the relationship between adropin and anthropological and psychological factors in schizophrenia. Nevertheless, to the best of our knowledge, no study has focused on adropin in schizophrenia based on human or animal models. If the disease characteristics could be explained through adropin, adropin might be a key target in the treatment of schizophrenia. Therefore, we measured adropin levels in patients with schizophrenia and performed a preliminary study to investigate adropin and the correlation with anthropological and psychological factors of this disease.

## Methods

### Participants

The study was performed in the Department of Psychosomatic Internal Medicine at Kagoshima University Hospital and Nishida Hospital (Kagoshima, Japan). Written informed consent was obtained from healthy volunteers (HV) and the patients with some schizophrenia spectrum and other psychotic disorders (SZ). We recruited participants using posters. All the participants who completed the experiments received 3,000 Japanese yen as compensation for their participation in the study.

In the schizophrenia spectrum and other psychotic disorders, according to the Diagnostic and Statistical Manual of Mental Disorders Fifth Edition,^14^ participants were required to be diagnosed with schizophrenia, schizoaffective disorder, or schizophreniform disorder. The diagnosis was made by psychiatrists.

In both groups, all participants were required to be between 45 and 65 years old, and participation had to be voluntary. Participants were excluded when psychiatrists assessed that participation in the study was inappropriate due to the severity of their psychotic symptoms.

### Study design

This was an observational case-control study aimed to investigate adropin levels and their correlations with various variables. The study protocol was approved by the Institutional Review Board of Kagoshima University Hospital (IRB approval number 230025 and 250090). Before the study, all participants were instructed to fast for 12 hours to obtain accurate laboratory test results. On the study day, in the morning, anthropological measurements (height, weight, and right and left handgrip strength (grip_R and grip_L)) were performed, followed by psychological tests and blood sample collection. Only in SZ, during psychological testing, the psychiatrists assessed the severity of their symptoms, followed by obtaining disease duration and the dosage of antipsychotic drugs converted to chlorpromazine equivalents from medical records.

### Psychological assessments

Participants were assessed for their psychological well-being using various questionnaires. First, for the evaluation of subjective depressed moods, anxiety, and trait emotional intelligence, we used the Zung Self-Rating Depression Scale (SDS),^15^ the State-Trait Anxiety Inventory Form X (STAI),^16^ which consisted of state and trait anxiety, and the Trait Emotional Intelligence Questionnaire-Short Form (TEIQue_SF),^17^ respectively. Second, for subjective sleep quality and perceived stress, the Pittsburgh Sleep Quality Index (PSQI)^18^ and the Perceived Stress Scale (PSS)^19^ were used, respectively. Finally, for apathy, we used the Japanese version of the Apathy Scale (AS),^20^ whose validity has been confirmed.^21^

### Psychiatric assessments

In SZ, the symptoms were assessed by psychiatrists using the Positive and Negative Syndrome Scale (PANSS).^22^ Briefly, PANSS is composed of 3 main domains: 7 items of positive symptoms, 7 items of negative symptoms, and 16 items of general psychopathology. Each item is scored from 1 to 7, as 1 indicates absence of symptoms and 7 indicates extreme symptoms. Therefore, the symptom severity ranges between 30 and 210.

### Laboratory tests

We obtained blood samples, and serum levels of calcium, copper, iron, selenium, zinc, vitamins A and B1, and ferritin were measured by a biochemical testing laboratory (Biken Co., Ltd., Kagoshima, Japan). Additionally, serum adropin levels were measured using ELISA kits of EK-032-35 (Phoenix Pharmaceuticals, USA), according to the manufacturer’s instructions.

### Software

We conducted the statistical analysis using Python version 3.11.5, pandas version 2.1.1, and Visual Studio Code version 1.97.2. Kendall’s correlation coefficient tau was calculated, and multiple comparison corrections with Bonferroni’s correction or false discovery rates were applied using statsmodels version 0.14.0. Data were visualized using seaborn version 0.13.2 and matplotlib version 3.8.0.

### Statistical analysis

First, we visually analyzed the variables by boxplots and estimated kernel density. Then, we divided the parameters into three domains: (1) anthropological parameters, (2) psychological parameters, and (3) laboratory test parameters. For each domain, assuming that all variables followed nonparametric distribution due to small sample sizes, medians were compared between HV and SZ using the Mann-Whitney U test with Bonferroni’s correction.

Third, we calculated Kendall’s correlation coefficient tau using multiple comparison corrections via false discovery rate (FDR). Because the aim was to detect potential correlations, the FDR threshold was set at 0.1. Finally, for the difference in tau between HV and SZ, each tau was subjected to an inverse hyperbolic tangent transformation to address boundedness and the difference was evaluated using a normal approximation-based Z-test with multiple comparison corrections via FDR. The FDR threshold was set at 0.05 because the aim was to compare the correlation between groups.

## Results

The study was performed from October 17, 2022, to October 2, 2023. A total of 20 participants (HV: 10; SZ: 10) completed the study, and no participants were excluded, though one participant in SZ had a comorbidity of diabetes mellitus with a stabilized blood sugar level. In SZ, all participants were hospitalized. The variables of study participants are summarized in the Supplementary Table.

For the PANSS, we only present general psychopathology scores, because it includes both positive and negative symptoms.^23^ PSQI, PSS, and grip_R indicated significant differences between groups. For the adropin levels, estimated kernel density and boxplots showed no significant differences (Supplementary Table 1; Fig 1). Additionally, paired correlations of all variables, along with their linear regression lines are displayed in the supplementary figure (Supplementary Fig S1).

**Figure 1.**
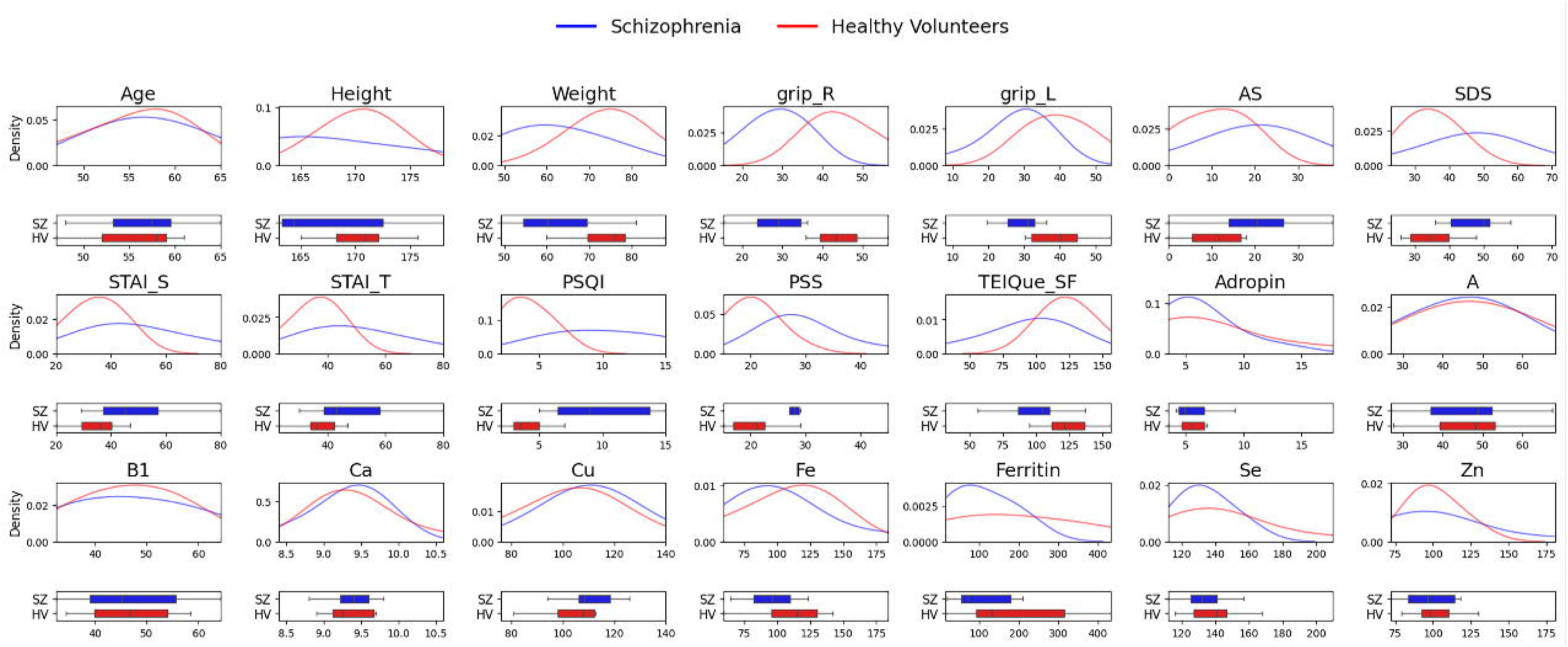
Estimated kernel densities and boxplots of the variables. AS = Apathy Scale, Ca = calcium, Cu = copper, Fe = iron, HV = the group of healthy volunteers, PANSS = Positive and Negative Syndrome Scale, PSQI = Pittsburgh Sleep Quality Index, PSS = Perceived Stress Scale, SDS = Zung Self-Rating Depression Scale, Se = selenium, STAI = State-Trait Anxiety Inventory Form X, STAI_S = state anxiety in STAI, STAI_T = trait anxiety in STAI, SZ = the group of patients with some schizophrenia spectrum and other psychotic disorders, TEIQue_SF = Trait Emotional Intelligence Questionnaire Short Form, Zn = zinc.

### Kendall’s correlation coefficients of tau

When the group was not divided, various significant correlations were observed, and most of them were related to psychological tests and anthropological parameters (Supplementary Fig S2).

In SZ, adropin and grip_R negatively correlated (τ = -0.82, *P* = 0.066), and selenium and state anxiety in the STAI (STAI_S) positively correlated (τ = 0.84, *P* = 0.066) (Fig 2). On the other hand, in the HV, only the positive correlation between grip_R and grip_L (τ = 0.82, *P* = 0.075) was indicated as significant (Fig 3).

**Figure 2.**
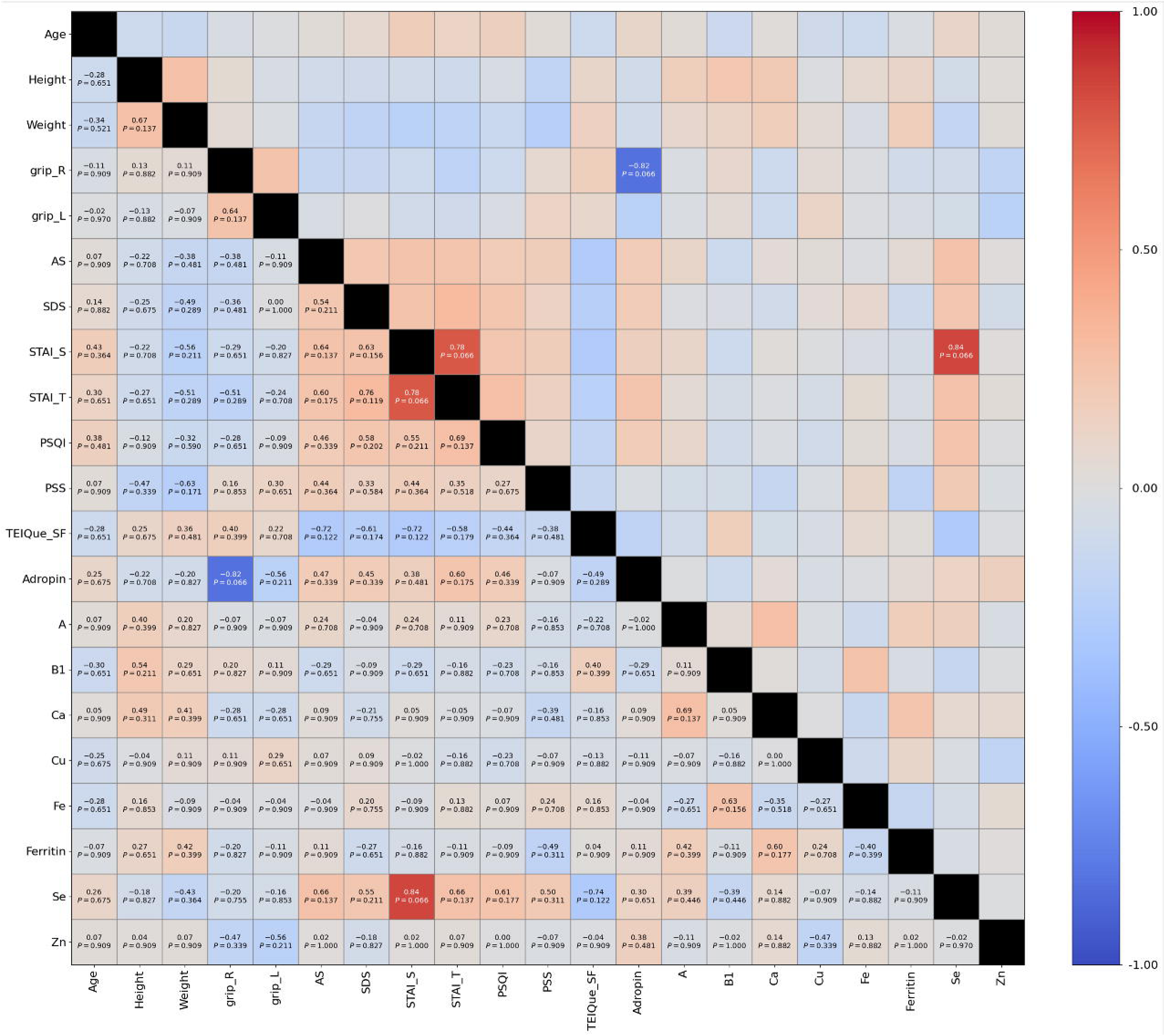
Correlation coefficients in participants with schizophrenia. AS = Apathy Scale, Ca = calcium, Cu = copper, Fe = iron, HV = the group of healthy volunteers, PANSS = Positive and Negative Syndrome Scale, PSQI = Pittsburgh Sleep Quality Index, PSS = Perceived Stress Scale, SDS = Zung Self-Rating Depression Scale, Se = selenium, STAI = State-Trait Anxiety Inventory Form X, STAI_S = state anxiety in STAI, STAI_T = trait anxiety in STAI, SZ = the group of patients with some schizophrenia spectrum and other psychotic disorders, TEIQue_SF = Trait Emotional Intelligence Questionnaire Short Form, Zn = zinc.

**Figure 3.**
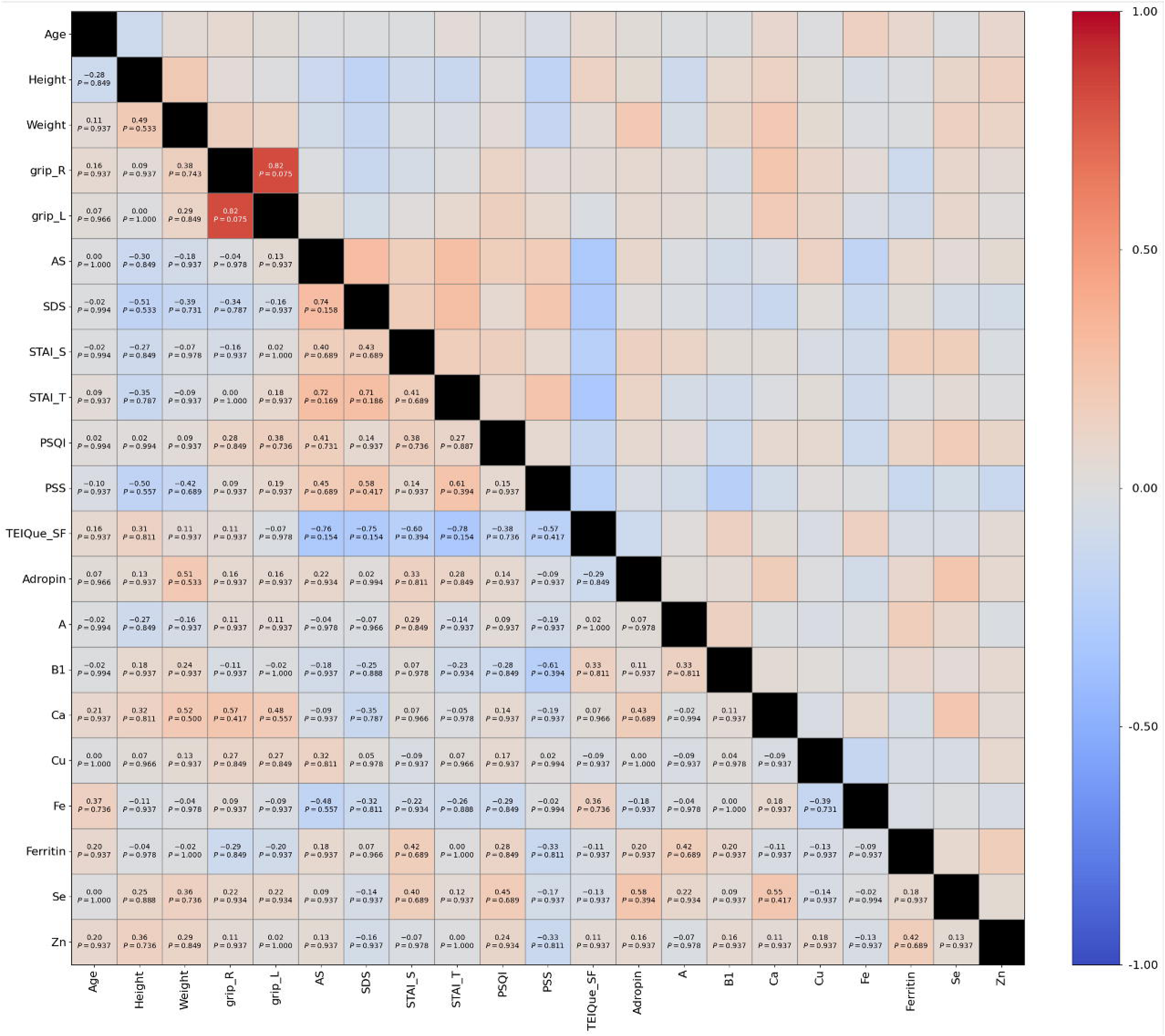
Correlation coefficients in healthy volunteer participants. AS = Apathy Scale, Ca = calcium, Cu = copper, Fe = iron, HV = the group of healthy volunteers, PANSS = Positive and Negative Syndrome Scale, PSQI = Pittsburgh Sleep Quality Index, PSS = Perceived Stress Scale, SDS = Zung Self-Rating Depression Scale, Se = selenium, STAI = State-Trait Anxiety Inventory Form X, STAI_S = state anxiety in STAI, STAI_T = trait anxiety in STAI, SZ = the group of patients with some schizophrenia spectrum and other psychotic disorders, TEIQue_SF = Trait Emotional Intelligence Questionnaire Short Form, Zn = zinc.

Differences in correlation coefficients for each variable are displayed in Fig 4. A significant negative difference was observed only in the correlation between adropin and grip_R (difference of τ = -0.98, *P* = 0.013). Significant findings in the further discussions are summarized in Table 1.

**Figure 4.**
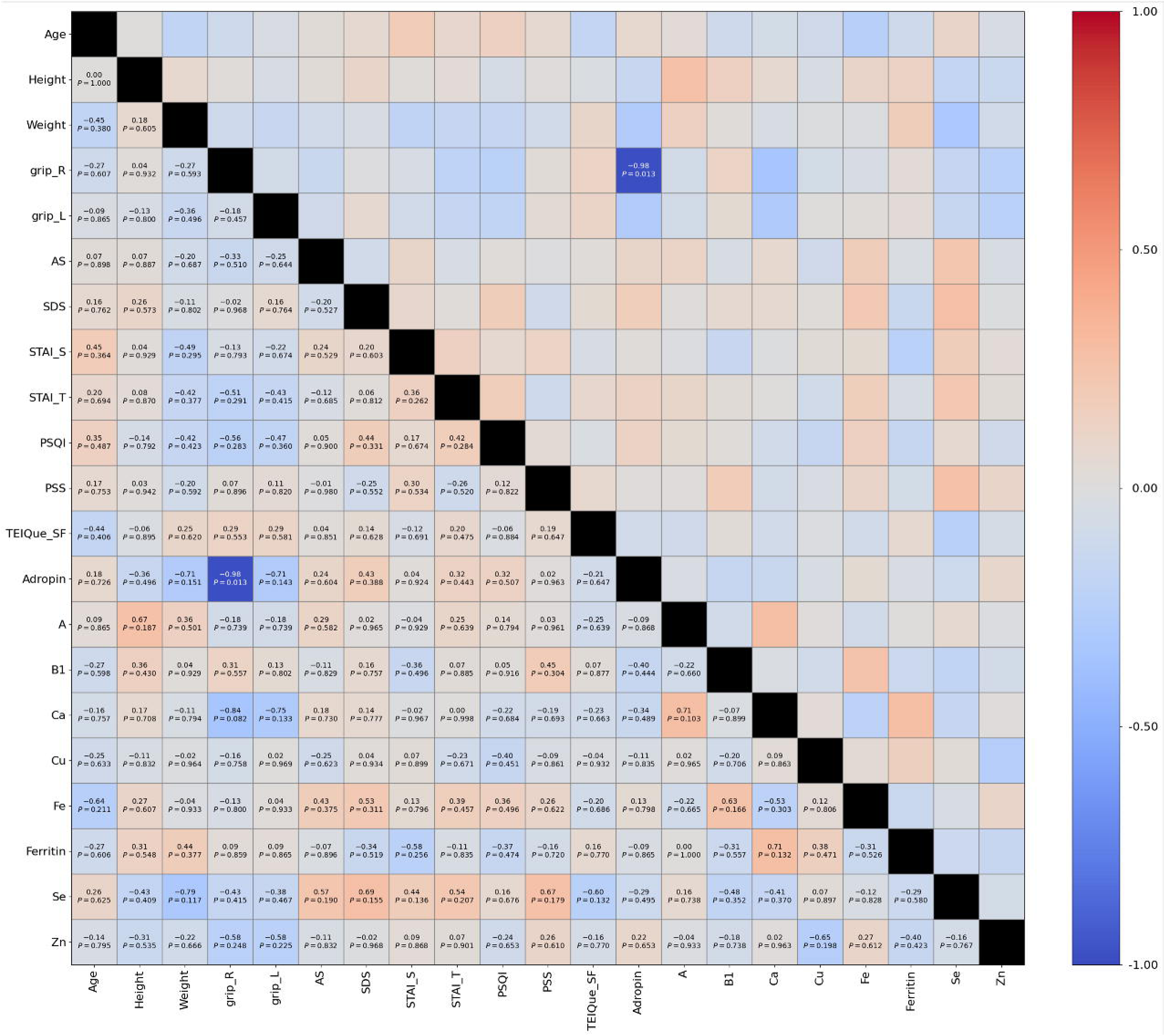
Correlation coefficients difference between groups. AS = Apathy Scale, Ca = calcium, Cu = copper, Fe = iron, HV = the group of healthy volunteers, PANSS = Positive and Negative Syndrome Scale, PSQI = Pittsburgh Sleep Quality Index, PSS = Perceived Stress Scale, SDS = Zung Self-Rating Depression Scale, Se = selenium, STAI = State-Trait Anxiety Inventory Form X, STAI_S = state anxiety in STAI, STAI_T = trait anxiety in STAI, SZ = the group of patients with some schizophrenia spectrum and other psychotic disorders, TEIQue_SF = Trait Emotional Intelligence Questionnaire Short Form, Zn = zinc.

**Table 1.** Significant correlations found in the analysis of Kendall’s correlations and their differences.

| Correlations | | Total ($\tau$, $P$) | HV ($\tau$, $P$) | SZ ($\tau$, $P$) | Difference ($\tau_{\text{diff}}$, $P$) |
| --- | --- | --- | --- | --- | --- |
| Adropin | Grip_R | $\tau = -0.14$, $P = 0.720$ | $\tau = 0.16$, $P = 0.937$ | **$\tau = -0.82$, $P = 0.066$** | **$\tau_{\text{diff}} = -0.98$, $P = 0.013$** |
| | Selenium | $\tau = \mathbf{0.44}, P = \mathbf{0.053}$ | $\tau = 0.58, P = 0.394$ | $\tau = 0.30, P = 0.651$ | $\tau_{\text{diff}} = -0.29, P = 0.495$ |
| Ferritin | PSS | $\tau = \mathbf{-0.44}, P = \mathbf{0.053}$ | $\tau = -0.33, P = 0.811$ | $\tau = -0.49, P = 0.311$ | $\tau_{\text{diff}} = -0.16, P = 0.720$ |
| Grip_L | Grip_R | $\tau = \mathbf{0.70}, P = \mathbf{0.001}$ | $\tau = \mathbf{0.82}, P = \mathbf{0.075}$ | $\tau = 0.64, P = 0.137$ | $\tau_{\text{diff}} = -0.18, P = 0.457$ |
| Selenium | STAI_S | $\tau = \mathbf{0.44}, P = \mathbf{0.053}$ | $\tau = 0.40, P = 0.689$ | $\tau = \mathbf{0.84}, P = \mathbf{0.066}$ | $\tau_{\text{diff}} = 0.44, P = 0.136$ |

The false discovery rate threshold was set at 0.05 in the correlation analysis and was set at 0.10 in the difference of correlations.
AS = Apathy Scale, HV = healthy volunteers, PSS = Perceived Stress Scale, STAI = State-Trait Anxiety Inventory Form X, STAI\_S = state anxiety in STAI, SZ = the group of patients with some schizophrenia spectrum and other psychotic disorders, $\tau_{\text{diff}}$ = the difference of $\tau$ calculated as the subtraction of tau in HV from SZ.

## Discussion

### Adropin levels in SZ are the same as in HV

The adropin levels are consistent with previous studies that measured adropin using the same ELISA kits,^24–27^ suggesting that the measurement was accurate and reliable. The levels were not significantly different between HV and SZ. Therefore, schizophrenia may not alter adropin levels.

### Interpretation of Kendall’s correlation test analyses

When the study group was not divided, the intergroup association in psychological tests was natural because psychological well-being involves various overlapping factors, such as anxiety, depressed moods, emotional intelligence, perceived stress, sleep quality, and vigor, which have intricate relationships. The intergroup association in anthropological parameters was also natural, reflecting fundamental proportional relationships in human morphology.

However, most of the associations were not detected when the group was divided. Although one possible reason is the reduced statistical power due to small sample size, the variables’ distributions might indicate another source of statistical bias, known as range restriction.^28^

In our study, psychological test scores, especially PSQI and PSS, were distributed differently between groups (Fig 1). On the contrary, laboratory tests, especially vitamins, selenium, and zinc, were distributed similarly. In anthropological parameters, handgrip strength and weight were distributed differently, but age and height were distributed relatively similarly. Thus, the difference in distributions may suggest that range restriction has influenced the detectability of associations. Based on these considerations, the correlation between selenium and adropin or STAI_S and ferritin and PSS might be important findings.

#### Statistical interpretations of significant correlations

As displayed in Table 1, significant correlations (i.e., correlations that showed statistical significance in the total group, in either group, or in the group difference, but were not attributed to natural or fundamental relationships) could be divided into three types: (1) not significant in the total group but significant in either SZ or HV, and the difference was statistically significant (adropin and grip_R), (2) not significant in both SZ and HV, with a non-statistically significant group difference, but significant in the total group (adropin and selenium, and ferritin and PSS), and (3) significant in the total group and either SZ or HV, but the difference is not significant (selenium and STAI_S, and grip_R and grip_L).

In the first type of correlation, a significant difference between groups was observed, and the significance was indicated in either group, which might reflect a real effect modulated by a group factor. In the second type of correlation, the non-detection of the correlation between ferritin and PSS in each group might be attributed to range restriction, considering the discrepant distributions of PSS in their boxplots (Fig 1). On the other hand, the distributions of adropin and selenium were similar across groups, suggesting that the lack of significance in each group may be due to insufficient statistical power rather than range restriction. Finally, in the third type of correlation, the observed correlation pattern of grip_R and grip_L differed from that of selenium and STAI_S. First, the correlation between grip_R and grip_L was relatively high in both groups, and the correlation in the total group was also relatively high. However, given that the scatter plot follows an approximately linear trend, as confirmed by the linear regression (Supplementary Figure), the non-detection of the correlation in SZ might be attributed to insufficient statistical power. Second, the correlation between selenium and STAI_S was high in SZ, but only significant in HV. Moreover, the scatter plot follows an approximately linear trend in SZ (Supplementary Figure). Therefore, the detected correlation in the total group might primarily reflect the strong correlation in SZ.

### Psychophysiological implications of the findings

Herein, we discuss the observed significant correlations from psychophysiological aspects, assuming the statistical validity discussed in the previous section.

#### Correlation between the right and left handgrip strength

The correlation between grip_R and grip_L is natural, but the correlation was not detected in SZ. In SZ, handgrip strength is lower than HV, aligning with previous research.^29,30^ Although the correlation between the right and left hand is not indicated in previous research, it can be inferred that the decrease in handgrip strength in both hands weakened the fundamental correlation observed in HV.

#### Correlation between the perceived stress and ferritin levels

The correlation between ferritin and PSS may suggest that perceived stress reduces stored iron or that reduced stored iron is associated with higher stress. Findings such as decreased intestinal ferritin absorption in rodents^31^ and lower ferritin levels in patients with coronavirus disease in intensive care units with psychiatric comorbidity^32^ align with our finding. Although the detected negative correlation between ferritin and PSS seems plausible based on the previous research, it should be noted that ferritin level is generally discussed in terms of iron deficiency.

#### Correlation between selenium and state anxiety

The positive correlation between selenium and state anxiety in SZ may suggest that selenium elevates anxiety levels in a state or that elevated selenium levels might be associated with a temporary high level of anxiety. The mechanism of selenium in the brain includes antioxidative effects induced by selenoproteins,^33,34^ neurotransmission,^35–37^ and neuroprotection.^38^

An association of low levels of selenium with schizophrenia has been reported,^39,40^ and the improvement of the schizophrenic symptoms by the co-supplementation of selenium and probiotics^41^ is consistent with the findings. In contrast, a meta-analysis indicated no significant associations of selenium levels with schizophrenia.^42^

In the present study, the correlation between state anxiety and selenium in SZ was not negative but positive. The normal selenium levels differ greatly between studies and countries,^43,44^ and a recent Japanese government study reported that the mean Japanese blood selenium levels were 180 μg/L.^45^ Therefore, the mean levels of 144.0 in HV can be considered as normal. However, assuming an optimal selenium range of around 80μg/L regarding anxiety or depression,^46,47^ adverse effects of selenium including transient anxiety^48^ might be plausible for the observed correlation in the present study.

#### Correlation between adropin and selenium

The role of adropin in the central nervous system is extensively described by Shahjouei et al., focusing on the signaling pathways.^49^ In schizophrenia, briefly, Akt/GSK3β pathway, which is the phosphorylation and inactivation of glycogen synthase kinase 3β (GSK3β) through the signaling molecule of Akt, is considered a key mechanism of antipsychotics targeting dopamine D2 receptors.^50^ Interestingly, the downregulation of GSK3β is reported in the condition of selenium deficiency.^51^ Moreover, adropin mediates the redox signaling through various pathways^52^ and selenium is a key factor in these pathways.^53^ Therefore, the association of adropin with selenium through GSK3β regulation and redox signaling appears plausible.

#### Correlation between adropin and handgrip strength in schizophrenia

The association of adropin with grip_R may reflect a real effect modulated by a group factor, according to the previous statistical discussion. Herein we discuss from the perspectives of (1) neural recognition molecule-3 (NB-3)/Notch 1 signaling pathways and (2) interaction with dopamine. First, in rodents, adropin-knockout mice decreased locomotor activities and ambulatory movements, which is attributed to the NB3/Notch1 signaling pathways^54^ functioning in both the developmental and post-developmental stages.^55^ However, the neuromuscular mechanism has not been focused, suggesting that the NB3/Notch 1 pathway’s contribution to the handgrip strength is inconclusive.

Second, an interaction between adropin and D2 receptors might be inferred since a potential adropin receptor G-protein coupled receptor 19 (GPR19)^56^ has a similar structure to the dopamine D2 receptors.^57^ In schizophrenia, a key mechanism is nigrostriatal pathways rather than mesolimbic pathways,^59^ patients exhibit overexpression of striatum D2 receptors, antipsychotics inhibit dopamine release, and the chronic antipsychotic treatment downregulates dopamine synthesis in some patients.^60,61^ Therefore, the comparable adropin levels in SZ to those of HV might be plausible if antipsychotics downregulated dopamine synthesis. On the other hand, if the dopamine levels were not comparable in SZ to those in HV, it can be hypothesized that adropin might be modulated by negative feedback for D2 dysregulation. Last but not least, these discussions should be carefully interpreted because we could not evaluate dopamine levels in the present study.

### Limitations

First, statistical results should be carefully interpreted by considering the small sample size, since this was a preliminary study and the main objective was to present the potential role of adropin in schizophrenia. However, we have discussed results carefully and consistently by considering the sample size shortage. Second, it should be noted that all participants in SZ were hospitalized. The hospital environment may introduce bias in handgrip strength or psychological well-being, but the patients performed physical activities and were not confined to bed. Third, because this was a preliminary study performed in men, the results should not apply to women. Last, the exclusion criteria did not include comorbidities. One participant in SZ had diabetes mellitus, which has extensively suggested the strong association between schizophrenia^62^ and adropin.^63^

## Conclusion

Upon considering the limitations, to the best of our knowledge, we first provide preliminary evidence of the association of adropin with schizophrenia. The present study provides a foundation for future studies and sheds light on adropin in schizophrenia, which might be a potential therapeutic target in schizophrenia.

## Data Availability

All data produced in the present study are available upon reasonable request to the authors

## Abbreviations

D2: dopamine D2 receptor
FDR: false discovery rate
GPR19: G-protein coupled receptor 19
grip_L: left handgrip strength
grip_R: right handgrip strength
GSK3β: glycogen synthase kinase 3β
HV: the group of healthy volunteers
NB-3: neural recognition molecule-3
PANSS: Positive and Negative Syndrome Scale
PSQI: Pittsburgh Sleep Quality Index
PSS: Perceived Stress Scale
STAI: State-Trait Anxiety Inventory Form X
STAI_S: state anxiety in STAI
SZ: the group of patients with some schizophrenia spectrum and other psychotic disorders

## Acknowledgements

We acknowledge the support of Dr. Haruki Iwai for his comments on the study results, and Dr. Kenichiro Sagiyama for his technical assistance.

