## Supplementary Table 1 for "Investigation of the correlation of adropin with anthropological and psychological factors in schizophrenia: preliminary evidence from a case-control study"

Supplementary Table 1. The variables obtained from study participants.

| Domain | Variables | Total | HV | SZ | P value |
| --- | --- | --- | --- | --- | --- |
| Anthropological | Age | 56.4 ± 1.1 | 55.8 ± 1.5 | 57.0 ± 1.7 | > 0.999 |
|  | Height (cm) | 169.2 ± 1.1 | 170.4 ± 1.0 | 167.9 ± 1.9 | > 0.999 |
|  | Weight (kg) | 68.4 ± 2.5 | 74.5 ± 2.5 | 62.4 ± 3.4 | 0.106 |
|  | Grip_R (kg) | 36.5 ± 2.4 | 44.5 ± 2.3 | 28.4 ± 2.1 | 0.001 |
|  | Grip_L (kg) | 33.7 ± 2.4 | 39.8 ± 2.6 | 27.6 ± 2.7 | 0.086 |
| Psychological | AS | 15.4 ± 2.3 | 10.5 ± 2.3 | 20.2 ± 3.4 | 0.262 |
|  | PSQI | 7.0 ± 1.0 | 4.0 ± 0.5 | 10.0 ± 1.2 | 0.007 |
|  | PSS | 24.9 ± 1.6 | 20.4 ± 1.3 | 29.3 ± 2.1 | 0.024 |
|  | SDS | 41.2 ± 2.7 | 34.9 ± 2.2 | 47.5 ± 4.1 | 0.162 |
|  | STAI_S | 41.9 ± 3.4 | 34.7 ± 2.7 | 49.1 ± 5.4 | 0.409 |
|  | STAI_T | 42.7 ± 3.1 | 36.2 ± 2.4 | 49.2 ± 5.0 | 0.374 |
|  | TEIQue_SF | 109.1 ± 6.4 | 123.8 ± 5.6 | 94.4 ± 9.6 | 0.063 |
| Laboratory tests | Adropin (ng/mL) | 6.7 ± 0.9 | 7.1 ± 1.5 | 6.3 ± 0.9 | > 0.999 |
|  | Ca (mg/dL) | 9.4 ± 0.1 | 9.4 ± 0.2 | 9.3 ± 0.1 | > 0.999 |
|  | Cu (μg/dL) | 107.7 ± 3.7 | 104.6 ± 5.5 | 110.7 ± 5.2 | > 0.999 |
|  | Fe (μg/dL) | 105.5 ± 7.0 | 109.2 ± 9.4 | 101.8 ± 10.6 | > 0.999 |
|  | Ferritin (ng/mL) | 149.0 ± 27.1 | 191.8 ± 46.7 | 106.1 ± 22.5 | > 0.999 |
|  | Se (μg/L) | 138.9 ± 5.0 | 144.0 ± 8.8 | 133.8 ± 4.7 | > 0.999 |
|  | Vitamin A (μg/dL) | 46.6 ± 2.7 | 47.3 ± 4.1 | 45.9 ± 3.8 | > 0.999 |
|  | Vitamin B1 (ng/mL) | 46.9 ± 2.2 | 46.8 ± 2.9 | 47.0 ± 3.6 | > 0.999 |
|  | Zn (μg/dL) | 102.2 ± 5.4 | 101.0 ± 4.9 | 103.3 ± 10.0 | > 0.999 |
| Psychiatric | Disease duration (yrs) | N/A | N/A | 25.7 ± 3.7 | N/A |
|  | PANSS | N/A | N/A | 72.3 ± 5.8 | N/A |
|  | Antipsychotic drug dosage (mg) | N/A | N/A | 720.4 ± 138.5 | N/A |

Variables with the statistically significant group difference are indicated in bold. N/A indicates that the P value or variables were not applicable. All variables are indicated mean ± standard error of the mean.

AS = Apathy Scale, Ca = calcium, Cu = copper, Fe = iron, HV = healthy volunteers, PANSS = general psychopathology score in Positive and Negative Syndrome Scale, PSQI = Pittsburgh Sleep Quality Index, PSS = Perceived Stress Scale, SDS = Self-Rating Depression Scale, Se = selenium, STAI_S = State-Trait Anxiety Inventory-State Anxiety Scale, STAI_T = State-Trait Anxiety Inventory-Trait Anxiety Scale, SZ = Schizophrenia, TEIQue_SF = Trait Emotional Intelligence Questionnaire Short Form, Zn = zinc.
