## Supplementary figures and images for "Investigation of the correlation of adropin with anthropological and psychological factors in schizophrenia: preliminary evidence from a case-control study"

### Supplementary Figure 1

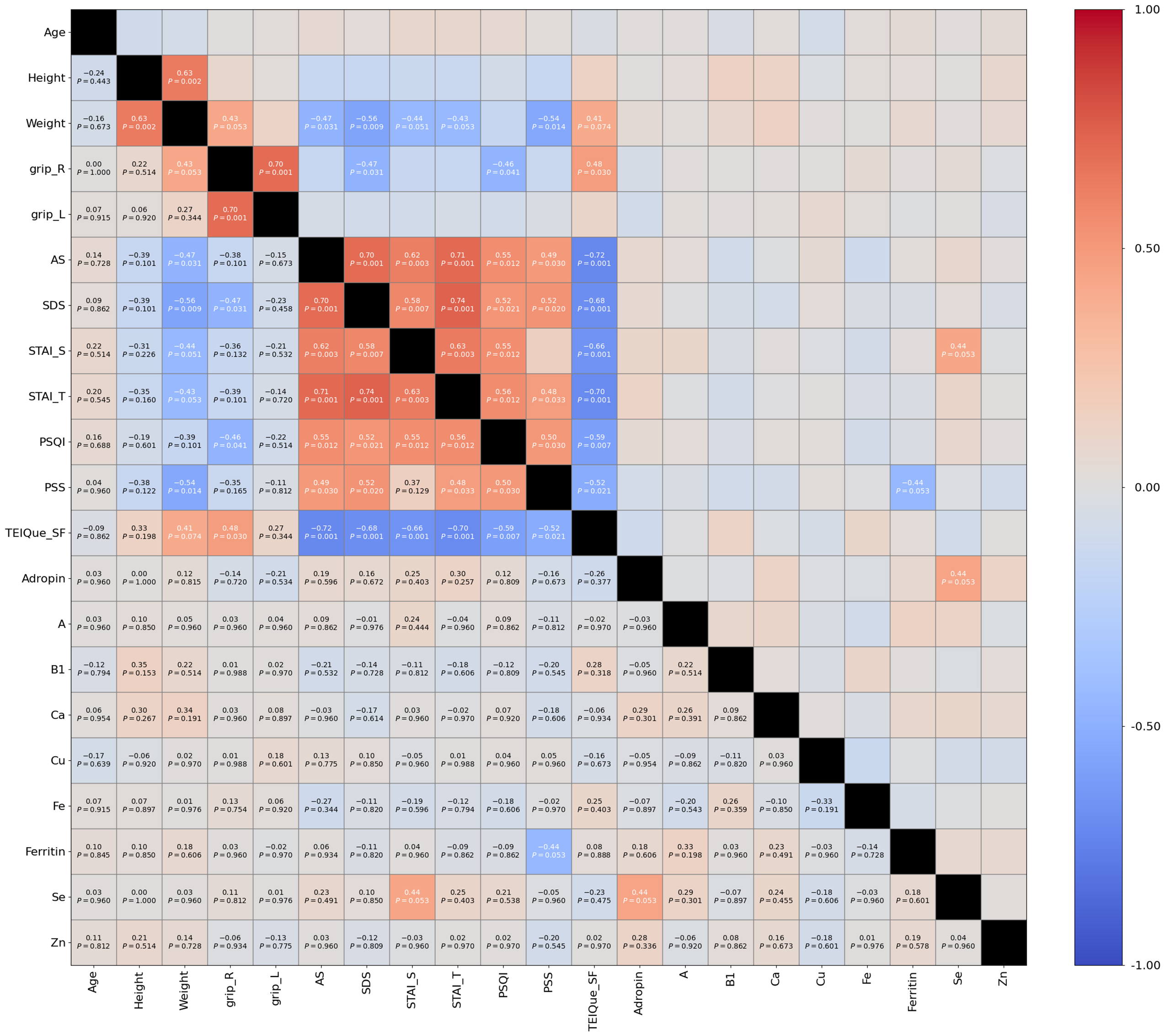

### Supplementary Figure 2

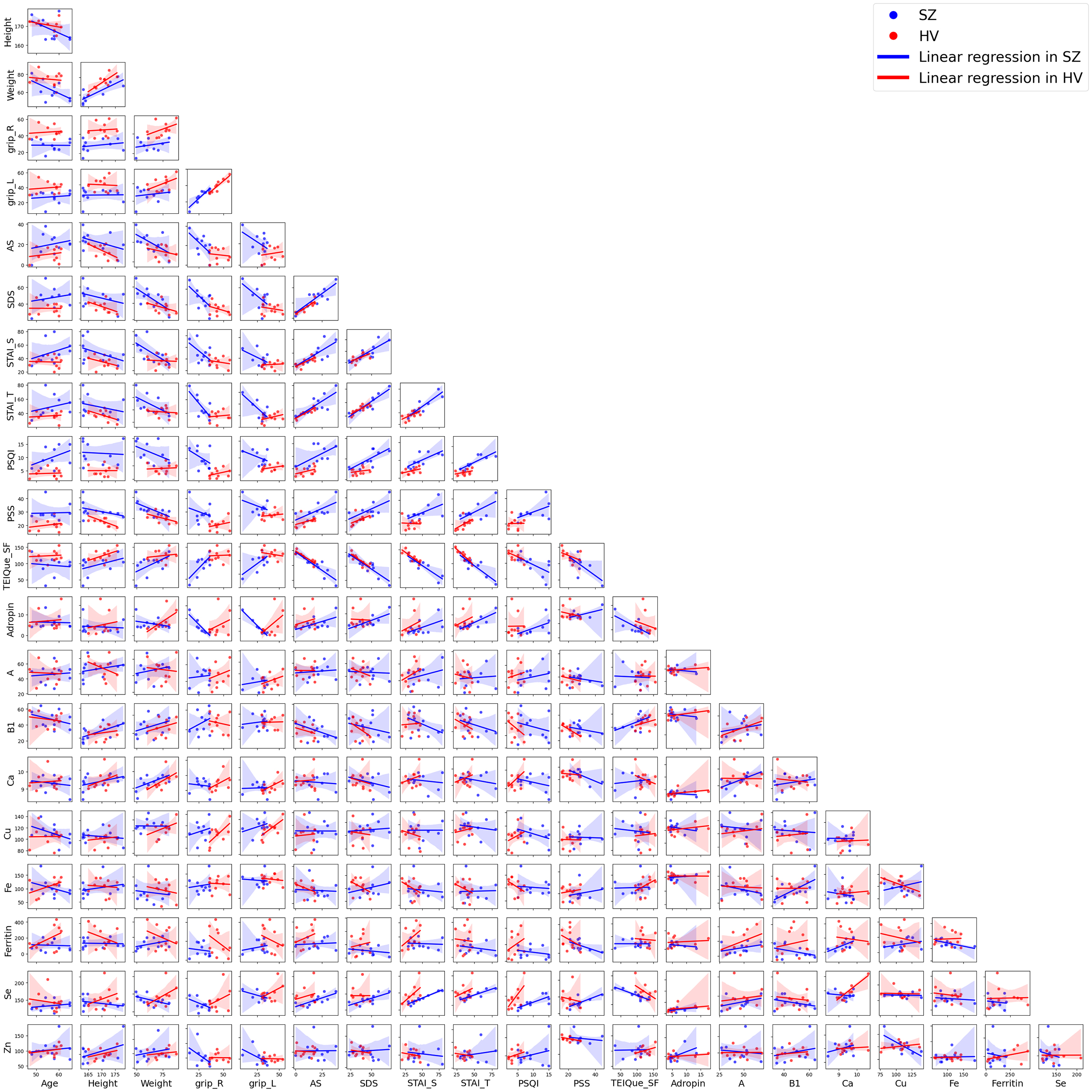
